# Utilising offspring genotype by proxy Mendelian randomization to investigate the causal effect of offspring traits on parental health

**DOI:** 10.1101/2025.06.11.25329457

**Authors:** Alesha A Hatton, Caroline Brito Nunes, Deborah Lawlor, David M. Evans

**Affiliations:** Institute for Molecular Bioscience, The University of Queensland, Brisbane, Australia; MRC Integrative Epidemiology Unit, University of Bristol, Bristol, United Kingdom; Population Health Science, Bristol Medical School, University of Bristol, Bristol, United Kingdom; Frazer Institute, The University of Queensland, Brisbane, Australia

## Abstract

Offspring can exert profound effects on the health of their parents. This is perhaps most apparent during the perinatal period, where the fetus influences processes that alter pre- and post-natal maternal physiology. In theory, it is possible to investigate the causal effect of offspring traits on parental health outcomes using Mendelian randomisation (MR), however, as parental and offspring genotypes are correlated, analyses need to be adjusted for the parent’s genotype to avoid confounding through the parental genome. Such analyses are difficult to perform at scale because of the paucity of cohorts across the world with large numbers of genotyped maternal- or paternal-offspring dyads and parent-offspring trios. In this manuscript, we explain how the causal effects of offspring traits on parental health outcomes can be investigated using Mendelian randomization (MR) and discuss the challenges in implementing such designs. We introduce the “offspring genotype by proxy” MR framework which can be employed in the absence of offspring genetic information to complement existing approaches in the triangulation of causal inference. The basic idea is to use parental genotypes to proxy the direct effect of their offspring’s genotype on their offspring’s own exposures. Specifically, we show how it is possible to proxy offspring genotype with paternal genotype when investigating causal effects of offspring traits on maternal health outcomes (and vice versa for paternal outcomes), which minimises the problem of confounding from the relevant parents’ genotype. We compare our framework to other MR designs that might be used to explore effects of offspring traits on parental health and investigate the consequences of model misspecification and spousal misclassification on statistical power and consistency. Given the increasing availability of datasets like the UK Biobank that (incidentally) include tens of thousands of genome-wide genotyped spousal pairs as well as large population based biobanks with linked health record data for first-degree relatives, we conclude that the offspring genotype by proxy MR approach could augment causal analyses of offspring exposures on their parents’ outcomes as implementation is not restricted to datasets with parent-offspring genotype information.

## Introduction

It is plausible that many offspring characteristics (e.g. behaviours, diseases, and the consequences of these) influence parental health. This is particularly likely to be the case for offspring perinatal traits and future maternal health, owing to the complex interplay between maternal and fetal physiological influences during the perinatal period (Haig, 1993). It has been suggested that fetal drive (genetic and/or environmental influences originating from the fetus) modifies maternal physiology as a potential mechanism to increase fetal nutrient delivery and optimise growth (Liu et al., 2014; Petry et al., 2007). For example, there is evidence that fetal genetic variants increase maternal glucose concentrations (Petry et al., 2017; Petry et al., 2011), and elevate the risk of gestational hypertension (Petry et al., 2014; Petry et al., 2016), preeclampsia (McGinnis et al., 2017) and hyperemesis gravidarum (Fejzo et al., 2024). Increased fetal growth (as indicated by gestational-age-adjusted birth weight and proxied by paternal transmitted birth weight scores) has been associated with a reduction in gestational duration and increased maternal systolic BP during pregnancy (Chen et al., 2020). Further, fetal genetic predisposition to a higher placental weight raises the risk of maternal preeclampsia and shortens gestational duration (Beaumont et al., 2023), potentially due to fetal insulin regulating placental weight. Fetal sex has also been associated with maternal increased fasting insulin sensitivity and secretion (Petry et al., 2022) and increased risk of pre-eclampsia and gestational diabetes in the presence of a male fetus (Broere-Brown et al., 2020). There are also proposed physiological changes in paternal health as a result of becoming a father. For example, first-time fathers exhibited a reduction in cortical volume and thickness of the precuneus when comparing structural magnetic resonance brain images of fathers who were scanned before and after their partners’ pregnancy with childless men at comparable time intervals (Paternina-Die et al., 2020). It has also been demonstrated that expectant fathers had lower testosterone and cortisol levels than controls (Berg & Wynne-Edwards, 2001; Gray et al., 2006). While such studies were observational in nature and were limited by sample size, these indications of potential physiological changes warrant further investigation.

Establishing whether observational associations are causal is important as this would support targeting offspring traits to improve parental health and family-based interventions. However, establishing causality in this context is particularly challenging. In part, this is due to the complex intergenerational interplay of genetic, environmental, and socio-economic factors that can confound the relationship between offspring traits and parental health outcomes. Additionally, observational associations can reflect reverse causality and bidirectional influences; for example, poor maternal health during pregnancy can affect the offspring’s birth outcomes, which in turn may further deteriorate maternal health postpartum and both maternal and paternal unhealthy behaviours pre-conception may influence the same behaviours in offspring and future paternal health.

In this manuscript, we explain how Mendelian randomization (MR; the use of genetic variants as unconfounded instrumental variables) can be used to investigate the causal effects of offspring traits on parental health outcomes. However, as genotyped maternal-(or paternal-) offspring pairs or parental-offspring trios are required to obtain unbiased estimate in this context, this poses a challenge in implementation. We introduce a new approach “offspring genotype by proxy” MR which can be employed in the absence of offspring genetic information to complement these existing methods in the triangulation of causal inference. This framework exploits studies, such as UK Biobank (UKB) that have information on tens of thousands of genome-wide genotyped spousal pairs. We also describe how this can be implemented using genotype data from unrelated individuals and linked health record data from their offspring and spouse. As described in more detail below, the method leverages the correlation between parental and offspring genotypes, using paternal (or maternal) genotypes to capture the direct effect of their offspring’s genotype on their offspring’s own exposure. We use causal graphs, asymptotic distribution theory and simulation studies to compare offspring genotype by proxy MR with existing approaches based on genotyped parent-offspring dyads and trios in terms of statistical power and (in)consistency due to horizontal pleiotropy. We also describe how offspring genotype by proxy MR can be implemented in both the one-sample and two sample MR frameworks and discuss potential studies for the large-scale application of our approach. In our explication, although we focus on the example of potential causal effects of offspring perinatal exposures (e.g. birth weight or small/large for gestational age) on their mother’s future post-natal health, analogously, this method can also be used to examine the causal effect of offspring perinatal traits on their father’s future health, and to explore potential effects of postnatal childhood and adult offspring exposures on both parents’ health.

### Mendelian randomization studies of the causal effect of offspring traits on parental health outcomes

MR is an epidemiological approach that uses genetic variants as instrumental variables to estimate the causal effect of environmental exposures on medically relevant outcomes and diseases (Smith & Ebrahim, 2003). MR approaches have advantages over traditional observational epidemiological methods in that they are robust to confounding, reverse causality and many types of bias. Whilst MR is a useful method for assessing causality, it relies on a number of assumptions which have previously been discussed at length (Evans & Davey Smith, 2015; Lawlor et al., 2017) (Figure 1a; supplementary methods). In theory it is possible to use offspring genetic variants to proxy offspring phenotypes and then use MR to estimate the causal effects of those offspring phenotypes on parental health outcomes. Using our illustrative example, offspring genetic variants associated with birth weight can be used to proxy fetal growth and these variants can be used in MR to investigate the potential causal effects of fetal growth (e.g. small/large for gestational age) on their mother’s future post-natal health outcomes. Such an approach would be subject to all the usual limitations inherent with MR studies, however, an additional issue specific to investigating the effect of offspring traits on maternal outcomes is the fact that maternal and offspring genotypes are correlated. Consequently, any association between the offspring genotype and maternal outcome may reflect confounding between the (correlated) maternal genotype and their own health outcome. This pathway violates the independence assumption of MR-that genetic variants are uncorrelated with factors that affect the outcome - and consequently may result in inconsistent estimates of the causal effect. Thus, it is important to block this pathway to provide consistent estimates of the causal effect of the offspring exposure on the maternal outcome. The most intuitive way of blocking this path is by conditioning on maternal genotype (i.e. at the same loci as in the offspring) by including it as a covariate in the MR analysis (as indicated in Figure 1b)(Lawlor et al., 2017). We refer to this approach herein as ‘MR with adjustment for maternal genotype’.

**Figure 1:**
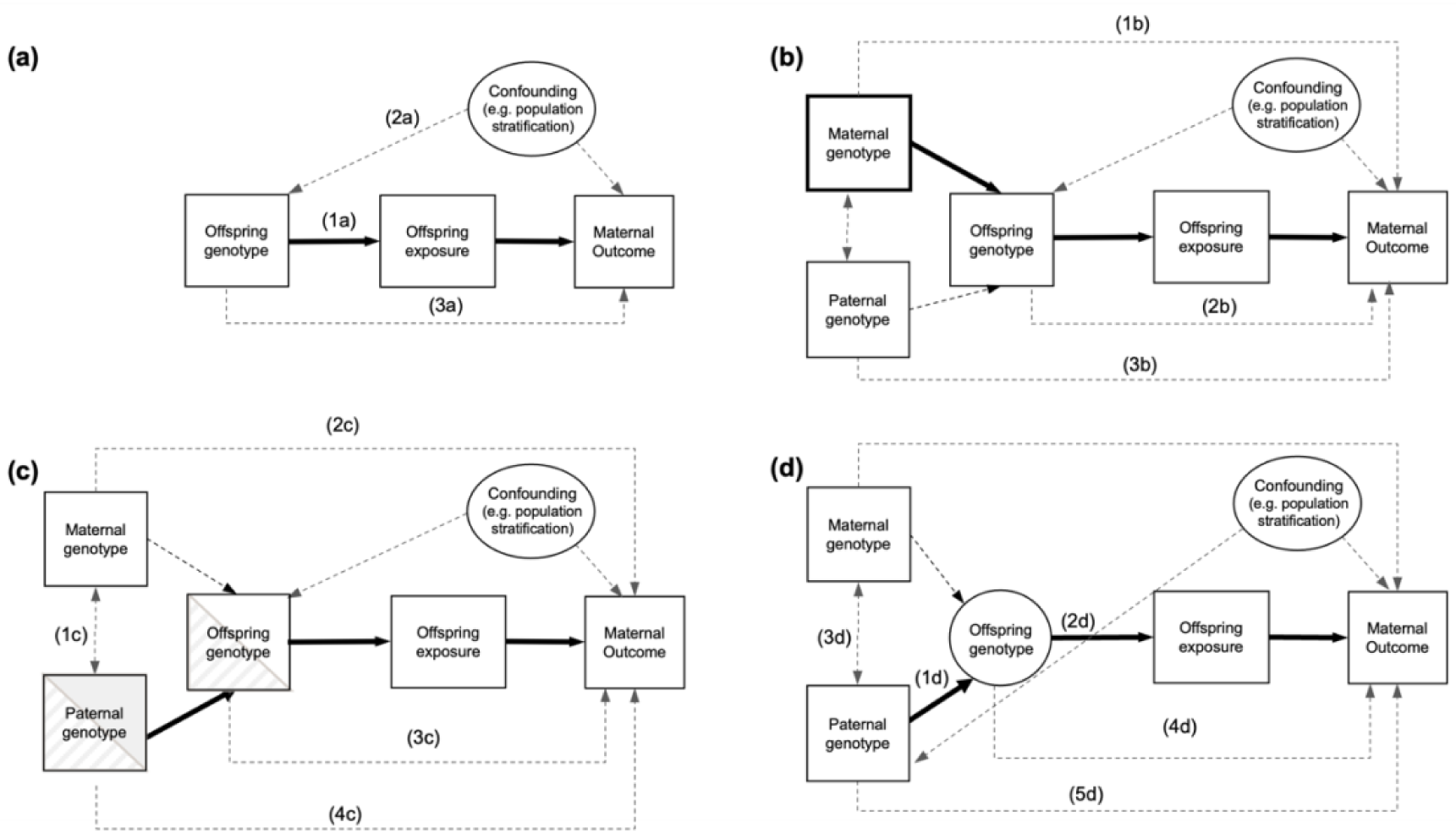
Causal diagram illustrating MR in the context of investigating the causal effect of an offspring exposure trait on a maternal health related outcome. Observed variables are represented by squares, whilst latent unobserved variables are represented by circles. Solid arrows indicate the paths of interest in MR. Dotted arrows indicate secondary paths or assumptions. Panel (a) illustrates the MR framework and its core assumptions of relevance (1a), independence (2a) and exclusion restriction (3a). Panel (b) illustrates how maternal genetic variants (that are correlated with the offspring genotype) may violate the independence assumption (1b). However, conditioning on maternal variants (as indicated by the bold box around maternal genotype) blocks path (1b). Path 2b illustrates how offspring genetic variants may violate the exclusion restriction assumption and how paternal genetic variants are a potential source of genetic confounding (3b). Panel (c) illustrates how utilising paternally transmitted alleles (which should be uncorrelated with maternal genotype in the absence of assortative mating; 1c) should protect MR analysis from confounding by the maternal genome (2c). Paths 3c and 4c indicate how paternally transmitted alleles in the offspring and father may result in a biased causal estimate. Panel (d) illustrates an offspring genotype by proxy MR approach where spousal information (i.e. father’s genotype) is utilized to proxy offspring genotype (1d), which in turn proxies the offspring trait (2d). Thus, paternal genetic variants can be used as instrumental variables to estimate the causal effect of an offspring exposure on a maternal outcome (as indicated by the path in bold). This obviates the need for offspring genotype (indicated by denoting offspring genotype as a latent unobserved variable in the figure). Under random mating, paternal and maternal genotypes should be uncorrelated (3d). The design also assumes the absence of pleiotropic paths from the offspring genome to the maternal phenotype (4d) and direct effects of the paternal genotype on maternal phenotype (5d).

Alternatively, the availability of genomic data on parent-offspring trios and/or mother-offspring dyads permits the identification of paternally transmitted alleles/haplotypes. Under random mating, proxying offspring phenotypes using paternally transmitted alleles (i.e. which should be uncorrelated with maternal genotype) should also protect MR analyses from confounding from the maternal genome (Figure 1c; referred to as ‘MR using paternally transmitted alleles’) (Zhang et al., 2015). As an example, Chen et al. constructed genetic scores using maternally transmitted, maternally untransmitted, and paternally transmitted alleles in 10,734 genotyped mother-offspring pairs to investigate a potential causal role of fetal growth on maternal physiological changes during pregnancy (Chen et al., 2020). The authors found the paternally transmitted birth weight score was associated with increased maternal systolic blood pressure during pregnancy and reduced gestational duration, suggesting a causal effect of offspring birthweight on these maternal phenotypes. Petry et al. similarly used 1,160 parent-offspring trios to infer parental transmitted and untransmitted alleles and identified an association between paternally transmitted fetal alleles at the *IGF2* locus and increased maternal glucose concentrations in the third trimester of pregnancy, again suggesting offspring effects on maternal physiology (Petry et al., 2011). In a similar fashion, identification of maternal transmitted haplotypes can be used to inform the likely maternal transmitted alleles and therefore inform the paternal transmitted alleles in the absence of paternal genotype (i.e. if only mother-offspring dyads are available). In theory, parentally transmitted haplotypes can be inferred without parental genomes (Hofmeister et al., 2024) using approaches designed to identify parent of origin effects. Paternally transmitted haplotypes can subsequently be used for MR provided linked health data is available for their parents.

Whilst good in theory, the problem with these approaches is that they are difficult to perform at scale because of the paucity of genotyped parent-offspring dyads and trios across the world’s cohorts (Evans et al., 2019). MR in general requires very large sample sizes in order to have adequate power to detect causal effects (Brion et al., 2012). In addition, the requirement to condition on the parental genotype to control for its role in confounding the offspring genetic instrument or performing analyses on haplotypes rather than genotypes will decrease statistical power to detect putative causal effects even further. The corollary is that currently many such analyses are likely to have low power to resolve causal effects. This is particularly true for effects of offspring traits on paternal health, with most cohorts having a smaller number of genotyped paternal-offspring pairs than they do maternal-offspring pairs. Consequently, this limits the utility and broader application of MR in investigating the causal effect of offspring traits on their parents’ health outcomes.

### Offspring genotype by proxy Mendelian randomization

In this section we introduce offspring genotype by proxy MR, an approach that in principle enables large scale MR studies of the causal effect of offspring traits on their parents’ outcomes when offspring genotypes are not available, but parental genotypes are. The basic idea is to utilise parental genetic variants to proxy offspring genotypes and consequently offspring exposures. For example, if the goal is in estimating the causal effect of an offspring exposure on some maternal outcome, then the IV would be genetic variants for the exposure in the offspring’s biological father. In theory, such an approach could be employed in combination with “traditional” MR analyses involving parent-offspring dyads (i.e. MR with adjustment for maternal genotype) or parent-offspring trios (i.e. MR using paternally transmitted alleles) and other informative designs to augment power and aid in the triangulation of causal evidence (Lawlor et al., 2016). We contrast these three approaches in Table 1, including their application in one- and two-sample MR and potential sources of bias, as discussed in greater detail subsequently in the manuscript.

**Table 1:** Comparison of three different MR approaches for estimating the causal effect of an offspring trait on a parental health outcome. We use the illustrative example of potential causal effects of offspring perinatal exposures on their mother’s future post-natal health to allow for explicit specification of the models. The three MR approaches include (1) Offspring genotype by proxy MR, (2) MR with adjustment for maternal genotype and (3) MR using paternally transmitted alleles.

| Method | Offspring genotype by proxy MR | MR with adjustment for maternal genotype | MR using paternally transmitted alleles |
| --- | --- | --- | --- |
| Description of the method (one sample MR) | Offspring genotype is proxied using paternal genotype at the genetic variant for the exposure in the father. MR is then performed using paternal genotype, offspring exposure and maternal health outcome as would be possible with genotyped spousal pairs and offspring phenotype information. | MR is performed using offspring genotype as the instrument, offspring exposure and maternal health outcome, with analyses adjusted for maternal genotype (at the same loci as in the offspring) as would be possible with genotyped mother-offspring pairs. | Paternal transmitted alleles are identified based on direct comparison of genotyped or local haplotype sharing using dense genome-wide trio data. MR is performed using the transmitted paternal alleles as the instrument, offspring exposure and maternal health. |
| Data required (one sample MR) | Genotyped fathers (or spousal pairs) with offspring exposure traits and maternal health outcomes. | Genotyped mother-offspring dyads with offspring exposure traits and maternal health outcomes. | Genotyped parent-offspring dyads or trios with offspring exposure traits and maternal health outcomes. |
| Description of the method (two sample MR) | SNP-exposure association estimates concerning the effect of offspring genotype on offspring exposure can be obtained from general GWAS summary statistics (assuming that there are no maternal/paternal effects at the locus).<br>SNP-outcome association needs to be obtained from the genotyped father under study i.e. the association between paternally proxied offspring genetic variants and the maternal outcome. | SNP-exposure association estimates concerning the effect of offspring genotype on offspring exposure.<br>SNP-outcome association estimates the effect of offspring genotype on maternal outcome (conditional on maternal genotype). | SNP-exposure association estimates concerning the effect of paternally transmitted alleles on offspring exposure.<br>SNP-outcome association estimates the effect of paternally transmitted alleles on maternal outcome. |
| Data required (two sample MR) | SNP-exposure sample requires own genotype and own phenotype. SNP-outcome sample requires genotyped spousal pairs or genotyped fathers with linked spousal phenotypic data. | SNP-exposure sample requires own genotype and own phenotype. SNP-outcome sample requires mother-offspring dyads. | Genotyped mother-offspring or father-offspring dyads or trios are required to identify paternally transmitted alleles for both the SNP-exposure and SNP-outcome associations. |
| Potential sources of bias | • Assortative mating on the exposure may introduce confounding through maternal genotype.<br>• Potential effects of paternal genetic variants on the maternal outcome (i.e. through paternal phenotypes).<br>• Horizontal pleiotropy through offspring genetic variants. | • Horizontal pleiotropy of offspring genetic variants.<br>• Confounding through the paternal genome | • Potential effects of paternal genetic variants (at paternally transmitted alleles) for the exposure on the maternal outcome (i.e. through paternal phenotypes).<br>• Horizontal pleiotropy of offspring genetic variants. |
| Limitations | Decreased power due to proxying offspring genotype. | Requires genotyped maternal-offspring dyads. | Requires genotyped parent-offspring dyads or trios. |

To understand why the offspring genotype by proxy approach would be informative, consider Figure 1d, which illustrates a credible way in which causal estimates of offspring traits on maternal outcomes can be obtained in the absence of offspring genotypes. As fathers transmit half their alleles to their offspring, it follows that paternal genotype will proxy offspring genotype, which in turn will proxy offspring phenotype (path 1d and 2d in Figure 1d). In the absence of horizontal pleiotropy between the paternal genotype and maternal outcome (and between offspring genotype and maternal outcome), this allows testing for a causal effect of the offspring trait on the maternal outcome. Using paternal genotype also obviates the requirement for individual-level genotyped mother-offspring pairs, although at the cost of a decrement in power compared to performing MR with adjustment for maternal genotype or MR using paternally transmitted alleles in genotyped mother-offspring pairs or parent-offspring trios (discussed in greater detail below). It is important to emphasise that if the goal is to estimate the effect of an offspring exposure on a maternal outcome, then in this design the instruments are paternal genetic variants. This is because the alternative i.e. using maternal genetic variants to proxy offspring genotype, would likely violate the independence assumption as maternal genetic variants influence offspring genotype and very plausibly maternal outcome. For example, if our aim was to explore the impact of offspring birthweight on future risk of maternal cardiovascular disease, using maternal genotype to proxy offspring genetic variants related to birthweight would also likely include variants that pleiotropically influence maternal blood pressure and glucose levels. A similar argument would also apply if one were to naively use paternal genetic variants to investigate the causal effect of offspring traits on paternal outcomes.

Given the increasing availability of datasets like the UK Biobank that (incidentally) include tens of thousands of genome-wide genotyped spousal pairs (Sudlow et al., 2015), the offspring genotype by proxy MR approach could augment causal analyses of offspring exposures on their parents’ outcomes. In theory, such an analyses can also be implemented in parous individuals even when genetic data is not available for the spouse (i.e. their offspring’s biological mother). Instead, large genetic datasets where data linkage to population-based registries and health records is available would permit implementation of the offspring genotype by proxy MR approach at a scale currently unattainable for MR approaches based on parent-offspring trios and/or mother-offspring dyads. For example, the Medical Birth Registry of Norway which comprises data on the mother, father and newborn for all births in the country after 16 weeks’ gestation can be linked to genotyped participants in HUNT (Brumpton et al., 2022) as well as national epidemiological registries which allow investigation of later life health outcomes (Irgens, 2000; Kvalvik et al., 2024). Data linkage of nation-wide, epidemiological registries for individuals as well as their parents, spouses, children, and siblings is also available in other countries e.g. FinRegistry (Viippola et al., 2023). The ability to link such registries to large genetic datasets containing (at least a subset of) the same individuals (e.g. The FinnGen study (https://www.finngen.fi/en) which contains genomic data with linkage to health register data of 500,000 Finnish biobank participants) allow for similar analyses to be conducted. Subsequently, large-scale investigation of causal effects of offspring traits on their parents’ health, such as that proposed here, could be conducted while only requiring genotype information for a single individual in each parent-offspring trio. In contrast, both MR conditional on maternal genotype and MR using paternally transmitted alleles, require genotyped mother-offspring or father-offspring dyads, even when implemented in the two-sample design (Table 1). As the offspring genotype by proxy MR approach relies on the assumption that any association between the paternally proxied offspring genetic variants for the exposure and maternal outcome operates through the offspring exposure, such analyses need to be performed in individuals that have not only had an offspring, but where the offspring exposure is present.

### Assumptions of offspring genotype by proxy MR

The offspring genotype by proxy MR approach involves the same three core assumptions as conventional MR analyses, however, under this approach there are additional nuances that need to be borne in mind (Figure 2). A detailed discussion of these assumptions is provided in the supplementary methods. The relevance assumption requires that the putative genetic instruments are robustly associated with the exposure of interest in the relevant population. In the offspring genotype by proxy approach, this depends upon the accuracy of spousal matching and on the statistical strength of association between offspring genotype and offspring exposure (Figure 2a). The independence assumption in MR requires no confounding between the genetic variants and outcomes of interest (Figure 2b). When investigating the potential causal effect of offspring traits on their mother’s (or father’s) health using MR this may be violated by confounding through the maternal (or paternal) genome. This is because maternal genotype will be correlated with offspring genotype (through transmission) and may also plausibly influence the maternal outcome. In the offspring genotype by proxy MR approach, we utilise the paternal genotype to proxy offspring genotype which should be uncorrelated with maternal genotype in the absence of assortative mating and therefore protect MR analyses from confounding by the maternal genome. This relies on the assumption of random mating for the exposure. Lastly, the exclusion restriction assumption stipulates the genetic instrument must only be associated with the outcome through the exposure. In the current context, it is therefore assumed that there is no directed path between paternal genetic variants and maternal outcome other than that going through the offspring exposure (Figure 2c). This includes the assumption of no pleiotropic paths from offspring genotype to maternal outcome (Figure 2c path 1c) and from paternal genotype to the maternal outcome through a paternal phenotype (Figure 2 path 2c).

**Figure 2:**
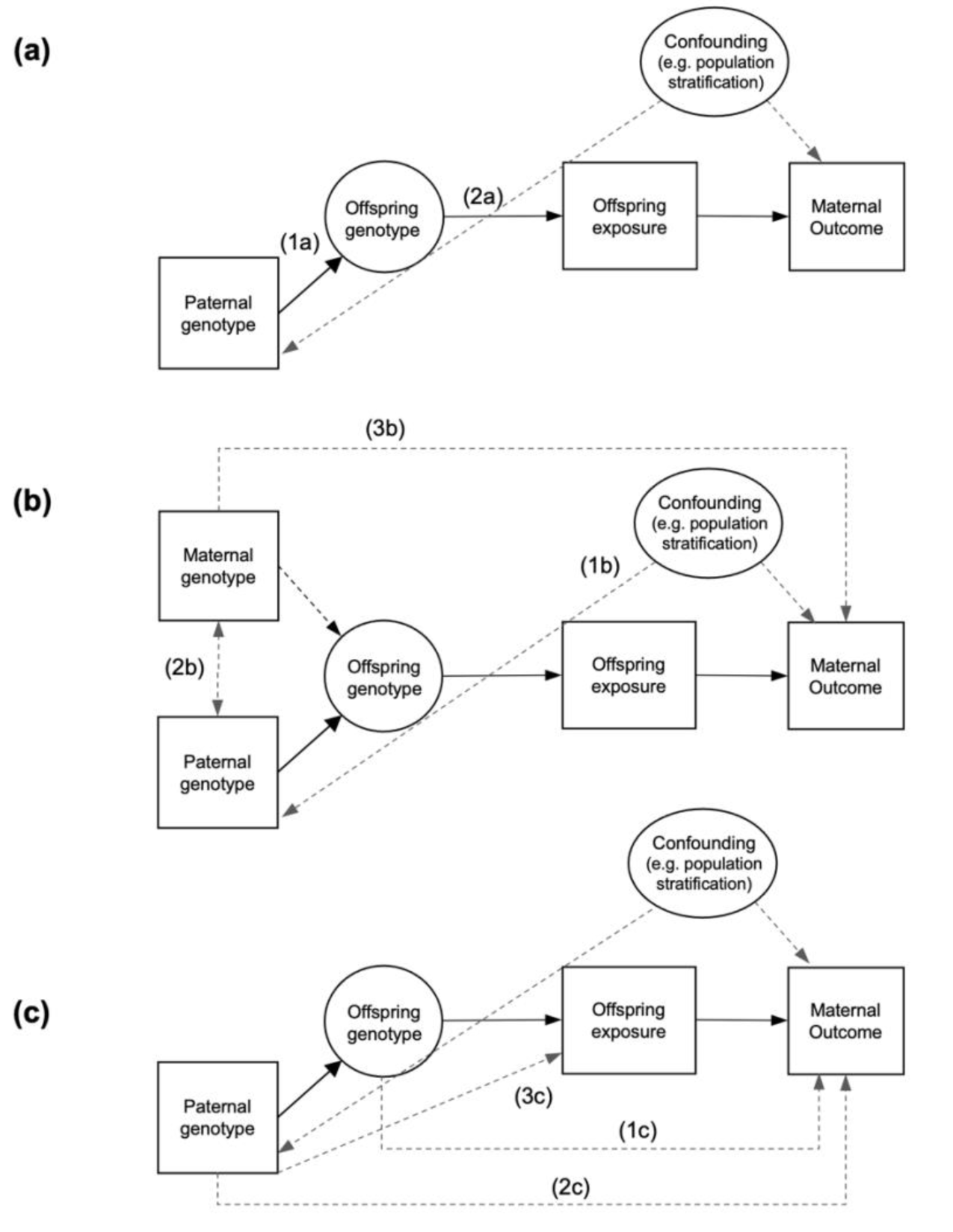
Causal diagram illustrating the offspring genotype by proxy MR approach and its underlying assumptions. Panel (a) illustrates the relevance assumption which requires correct matching of spousal pairs and for paternal genotype to be from the biological father of the offspring (path 1a) and the proxied offspring genetic variants to be statistically robustly associated with the exposure in the relevant population (path 2a). Panel (b) highlights the independence assumption whereby there are no confounders of the paternal genotype – maternal outcome relationship (1b). Further, genetic variants for the exposure in the father should not be associated with genetic variants in the mother (e.g. as might be the case under assortative mating on the exposure; path 2b) as this may create a path to maternal outcome if correlated variants in the mother have effects on her outcome (path 3b). Part (c) illustrates the exclusion restriction assumption which requires that no causal path exists from the paternal genotype (which is used to proxy the offspring genotype) to the maternal outcome other than through the offspring exposure (paths 1c and 2c). While dynastic effects from paternal genotype to offspring exposure (path 3c) do not violate the exclusion restriction assumption, these must be accounted for when estimating the causal effect.

### Exploring violation of the MR assumptions

#### Inconsistency of instrumental variable estimators under pleiotropy

We explore the consequences of violation of the exclusion restriction assumption by way of horizontal pleiotropy through either the offspring or paternal genome. Following asymptotic theory , we derived the large sample properties of the IV estimator for each of the three different MR approaches for estimating the causal effect of an offspring exposure on a maternal outcome: (1) Offspring genotype by proxy MR (Figure 1d), (2) MR with adjustment for maternal genotype (Figure 1b) and (3) MR using paternally transmitted alleles (Figure 1c). Derivations are provided in the supplementary material. For all derivations, we assume absence of assortative mating and confounding from maternal genotype (noting that in the absence of assortative mating all three approaches are robust to confounding from the maternal genotype at any level of association between maternal genotype and maternal outcome).

All three MR approaches yield inconsistent IV estimates when the assumption of no effect of offspring and/or paternal genetic effects on the maternal outcome is violated (i.e. horizontal pleiotropy). In the presence of horizontal pleiotropy through the offspring genotype, the probability limit of the IV estimator differed from the population level causal effect of offspring exposure on maternal outcome. For all three approaches this is equal to the effect of offspring genotype on maternal outcome divided by the effect of offspring genetic variant on offspring exposure. However, in the presence of horizonal pleiotropy through the paternal genotype, the probability limit of the IV estimator differed between the three approaches. The magnitude of difference between the probability limit of the IV estimator and the population level causal effect was largest for the offspring genotype by proxy MR design, equal to twice the effect of paternal genotype on maternal outcome divided by the effect of offspring genetic variant on offspring exposure. The was reduced by a factor of ½ for MR using paternally transmitted alleles and by a factor of ¼ for MR with adjustment for maternal genotype.

#### Misclassification of spousal pairs

Matching of spousal pairs based on demographic information, such as in the UK Biobank, is subject to misclassification. Further, correct matching of spousal pairs does not necessitate that both individuals are the biological parents of any reported offspring. In this section, we considered the consequences of (accidental) inclusion of incorrectly matched spousal pairs (or correctly matched spousal pairs that are not biological parents) on bias and statistical power. We performed a simple simulation study to identify the likely effect of such misclassification varying the proportion of randomly misclassified spouses (0%, 10%, 20%, 30% and 40% of spousal pairs), utilising the illustrative example of potential causal effects of offspring perinatal exposures on maternal post-natal health. Within this model we investigated two misclassification scenarios; (1) A male who is genetically unrelated to the offspring under consideration is incorrectly used to the proxy offspring exposure; (2) A genetically unrelated female is incorrectly paired with the father and his offspring. Full details for the data generation and simulation study can be found in the supplementary material and full simulation results in supplementary table 1.

The results from selected simulations are presented in Table 2. The effect of spousal pair misclassification depends on the true relationship between offspring and putative parents (assuming the exposure is reported by the true parent). In the first scenario, misclassification was associated with decreased strength of association between the instrument and the exposure, however, causal estimates were largely unbiased if instruments were strong. In scenario 2, the causal effect attenuated towards the null as misclassification increased. In this scenario instrument strength was not impacted as misclassification increased. In both scenarios, statistical power decreased as instrument strength decreased, and spousal pair misclassification increased.

**Table 2:** The impact of spousal pair misclassification on the instrumental variables estimate of offspring exposure on maternal outcome using offspring genotype by proxy MR.*

| Scenario 1: Male not related to offspring |  |  |  |  |  |  |
| --- | --- | --- | --- | --- | --- | --- |
| $V_q$ | Proportion misclassified | BetaIV(median) | SE(BetaIV) | Power | Fstat | Coverage |
| 0.5% | 0 | 0.198 | 0.12 | 0.41 | 62 | 0.96 |
| 0.5% | 0.1 | 0.196 | 0.13 | 0.34 | 50 | 0.95 |
| 0.5% | 0.2 | 0.197 | 0.15 | 0.31 | 40 | 0.96 |
| 0.5% | 0.3 | 0.191 | 0.17 | 0.22 | 30 | 0.96 |
| 0.5% | 0.4 | 0.193 | 0.20 | 0.20 | 23 | 0.97 |
| 2% | 0 | 0.199 | 0.06 | 0.91 | 251 | 0.96 |
| 2% | 0.1 | 0.198 | 0.07 | 0.82 | 202 | 0.94 |
| 2% | 0.2 | 0.198 | 0.07 | 0.75 | 160 | 0.96 |
| 2% | 0.3 | 0.196 | 0.08 | 0.65 | 122 | 0.95 |
| 2% | 0.4 | 0.196 | 0.10 | 0.51 | 91 | 0.96 |
| Scenario 2: Female not related to offspring |  |  |  |  |  |  |
| $V_q$ | Proportion misclassified | BetaIV(median) | SE(BetaIV) | Power | Fstat | Coverage |
| 0.5% | 0 | 0.198 | 0.12 | 0.41 | 62 | 0.96 |
| 0.5% | 0.1 | 0.176 | 0.12 | 0.33 | 62 | 0.95 |
| 0.5% | 0.2 | 0.156 | 0.12 | 0.29 | 62 | 0.97 |
| 0.5% | 0.3 | 0.135 | 0.12 | 0.21 | 62 | 0.94 |
| 0.5% | 0.4 | 0.115 | 0.12 | 0.17 | 63 | 0.93 |
| 2% | 0 | 0.199 | 0.06 | 0.91 | 251 | 0.96 |
| 2% | 0.1 | 0.178 | 0.06 | 0.81 | 250 | 0.93 |
| 2% | 0.2 | 0.158 | 0.06 | 0.75 | 250 | 0.91 |
| 2% | 0.3 | 0.137 | 0.06 | 0.63 | 250 | 0.84 |
| 2% | 0.4 | 0.118 | 0.06 | 0.49 | 252 | 0.75 |

\*Data simulations were based on 50,000 parent offspring trios across 1,000 replicates. A causal effect of $\beta_{XY} = 0.2$ was used i.e. the difference in mean maternal outcome in standard deviation units per unit increase in offspring exposure. The strength of confounding between the offspring exposure and maternal outcome was set at $\beta_{UX} = \beta_{UY} = 0.5$ . $V_q$ = variance explained by the offspring genetic variant in the offspring exposure. In scenario (1) a male who is genetically unrelated to the offspring under consideration is incorrectly used to the proxy offspring exposure, while in scenario (2) a genetically unrelated female is incorrectly paired with the father and his offspring.

### Application of Offspring genotype by proxy MR within the gene-by-environment MR framework

Certain gene-by-environment (GxE) interactions can be leveraged in MR studies by providing evidence for violation of the exclusion restriction assumption and in some situations, enabling estimation of the causal effect of the exposure on the outcome correcting for horizontal pleiotropy (Chen et al., 2008; Davey Smith, 2011; Spiller et al., 2019). In principle, the strategy requires an environmental variable that can be used to stratify the sample into two (or more) groups with different levels of the exposure, including one where the exposure is absent (the “no relevance” group), and consequently, where there should not be any within group association with the genetic instrument. Thus, any causal effect should only be observed in strata where the exposure manifests, but not in strata where it does not (similar to the situation in a negative control analysis). Indeed, the presence of a (spurious) “causal” association between the genetic instrument and the outcome in the no-relevance group can be used to quantify the degree of horizontal pleiotropy present in the analysis, and to subsequently correct any causal estimates for pleiotropy in the other groups. An important consideration when using this approach is that the instrument should not be associated with the phenotype used to stratify the sample as this may induce collider bias (Coscia et al., 2022).

The GxE MR approach is particularly relevant to the framework espoused in this manuscript in that spousal pairs can be stratified by parity (i.e. the presence or absence of an offspring shared between the spouses). Here nulliparous spousal pairs effectively act as negative controls, allowing for the identification of heterogeneity in effect estimates owing to non-causal influences. For example, if the offspring exposure is causal for the maternal health outcome, then genetic instruments for the exposure in the father should only be associated with maternal outcomes in spousal pairs whom have shared an offspring. Conversely, the “offspring” exposure could not plausibly influence the outcome in nulliparous pairs. Therefore, any associations between paternal genotype and maternal outcome in spousal pairs without an offspring would indicate that the apparent causal effect is spurious, potentially due to horizontal pleiotropy. Interestingly, utilizing GxE interactions in nulliparous spousal pairs is only possible when proxying offspring genotype using parental genotypes. It is impossible in MR with adjustment for maternal genotype and MR using paternally transmitted alleles which involve mother-offspring/father-offspring dyads and parent-offspring trios (which by definition involve parous mothers).

Finally, we note that there may be opportunities afforded by broken partnerships in modelling causal effects where these data are available (and conversely interpretational complexities where they are not). For example, consider a parent-offspring trio where the husband is biologically related to the child, but as he is now on his second marriage, his spouse is not. Assume also, that this couple have not had any biological children of their own. It is conceivable that the father’s offspring exerts some sort of causal effect on his spouse’s health, however, any effect will need to be mediated via post-natal pathways. In other words, this trio would form part of a no-relevance group if the focus was on estimating the long-term causal effect of perinatal offspring traits on maternal health. Similar designs have been proposed using adopted versus biological relatives (Hwang et al., 2022).

### Power of the offspring genotype by proxy MR approach

Genetic variants typically only explain small amounts of variation in exposure variables. Consequently, MR studies often need very large sample sizes in order to detect precise causal effects with appreciable power (Brion et al., 2012).This limitation is exacerbated in the offspring genotype by proxy MR approach as genetic variants in one of the parents are used to proxy offspring genetic variants, which in turn proxy the offspring exposure. As parental and offspring genotypes are correlated 0.5, the proportion of variation in the offspring exposure explained by the parental genotype will be reduced by a factor of 0.25 relative to that explained by the offspring’s own genotype. We investigated the power of three different approaches: (1) Offspring genotype by proxy MR, (2) MR with adjustment for maternal genotype and (3) MR using paternally transmitted alleles. Methods for the data simulations are outlined in the supplementary material. We also provide the estimated sample size for MR using the offspring genotype by proxy approach calculated from asymptotic theory (Brion et al., 2012) for varying proportions of variation in the offspring exposure variable explained by the offspring genetic instruments and confirm similar estimates were obtained to those estimated using simulations (supplementary table 2).

Figure 3 displays illustrative examples of the power to detect a causal effect using each of the three study designs assuming a true non-null causal effect of β_XY_ = 0.1. Here we report the minimum sample size required to obtain 80% power (type I error rate α = 0.05). For the offspring genotype by proxy MR approach, the variance explained by the genetic instruments is reduced by a factor of 1/4 relative to that of the mother-offspring pairs approach. Subsequently, our approach will be less powerful than similar-sized samples of genotyped mother-offspring pairs. Likewise, offspring genotype by proxy MR is less powerful than MR using paternally transmitted haplotypes. The corollary is that, in order to have enough power to resolve causal effects on its own given current sample sizes, offspring genotype by proxy MR would need to be employed in very large samples of spousal pairs using allelic scores that explain considerable proportions of variance in the offspring exposure variable. Despite this, as there is a paucity of large cohorts of genotyped mother–offspring pairs and parent-offspring trios around the world, the method could provide a complementary approach to causal inference owing to the availability of large numbers of genome-wide genotyped spousal pairs in publicly available genetic datasets such as the UK Biobank as well as large population based biobanks with linked health record data for first-degree relatives.

**Figure 3:**
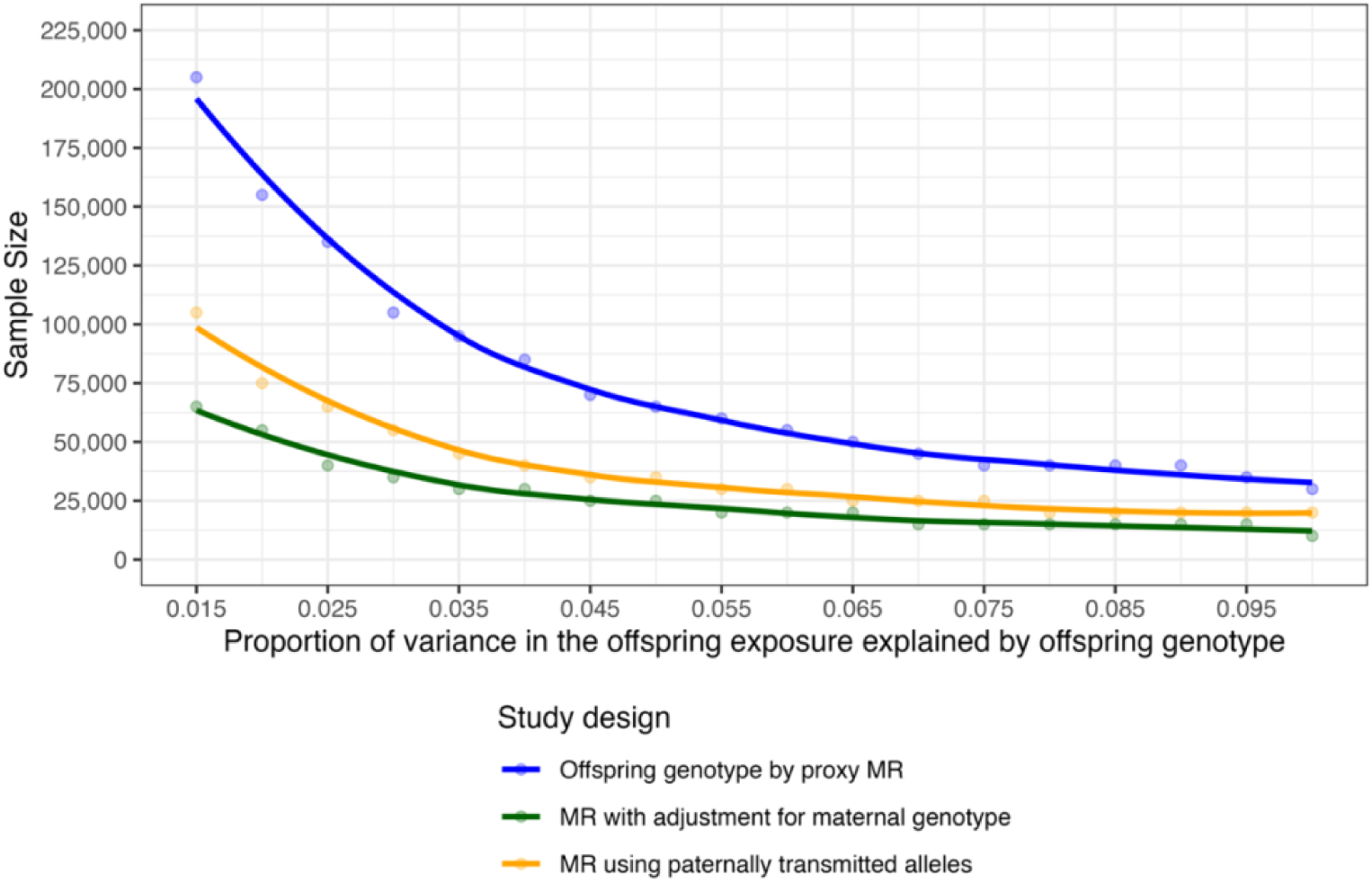
Illustrative examples of the sample sizes required for 80% power to detect a causal effect estimate of offspring exposure on maternal outcome using three MR approaches. The approaches were (1) Offspring genotype by proxy MR, (2) MR with adjustment for maternal genotype and (3) MR using paternally transmitted alleles. The sample size is presented for varying proportions of variance in the offspring exposure explained by the offspring genetic instrument. For each approach, a single observational unit represents a genotyped parent-offspring trio (even if not all are needed for the analysis). Power calculations were performed assuming a Type 1 error rate of α = 0.05 and a causal effect size of β_XY_ = 0.1. The causal effect is the difference in mean maternal outcome in standard deviation units per standard deviation increase in offspring exposure. The strength of confounding between the offspring exposure and maternal outcome was set at β_UX_ = β_UY_ = 0.5 and estimates were based on the median of 1,000 simulation replicates.

### Estimation of causal effects: one-vs two-sample MR

In one sample MR, the genetic variant, exposure and outcome are all measured in the same group of individuals and a causal estimate is calculated using e.g. two stage least squares. In contrast, in two sample MR the exposure and the outcome are measured in independent samples. Independent estimates of the SNP-exposure association and the SNP-outcome association are then combined to estimate the causal effect of the exposure on the outcome (Abraham, 1940; Pierce & Burgess, 2013). This is commonly done using the Wald Estimator. The advantage of the two sample approach is that it can utilise publicly available GWAS summary results statistics that are based on potentially hundreds of thousands of individuals, and therefore dramatically increase statistical power compared to the one sample situation (Hemani et al., 2018; Lawlor, 2016). In the offspring genotype by proxy MR design, it is essential to match the genetic variants for the offspring exposure in one parent with the outcome measured in the other. The most intuitive way of estimating the causal effect would be using two-stage least squares in the one-sample MR context, where in the first stage, the offspring exposure is predicted using the proxied offspring genotype i.e. using paternal (or maternal) genotype. The causal effect estimate is then obtained by regressing the maternal (or paternal) outcome on the predicted offspring exposure in the second stage of the analysis and calculating appropriate standard errors.

However, a causal effect can also be calculated using the Wald estimator using summary level data from different samples from the same underlying population. That is, estimates of the SNP-exposure association (i.e. between paternal genetic variants and the offspring exposure) and estimates of the SNP-outcome association (between paternal genetic variants and the corresponding maternal outcome) do not need to be taken from the same sample. The critical element is that estimates of the paternal genetic variant-maternal outcome association are obtained using correctly matched parents (i.e. biological parents) of the offspring. We present a diagram illustrating estimation of the causal effect of an offspring’s trait on their parent’s (maternal) health outcomes in Figure 4. β_zx_ represents the effect of offspring genotype on offspring exposure and β_xy_ the causal effect of offspring exposure on maternal outcome.

**Figure 4:**
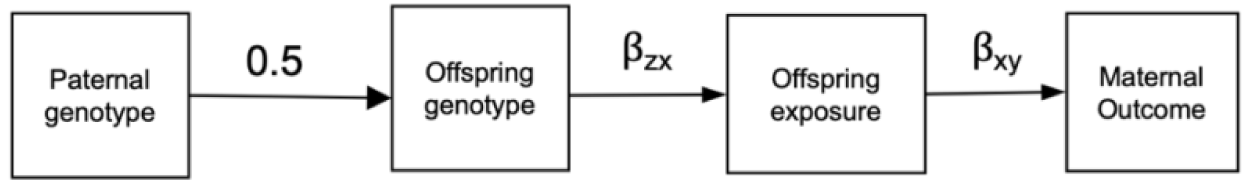
Estimating the causal effect of an offspring exposure on a maternal health outcome using offspring genotype by proxy MR. β_zx_ represents the effect of offspring genotype on the offspring exposure and β_xy_ is the causal effect of offspring exposure on the maternal outcome. In one-sample MR, the genetic instrument, exposure and outcome are measured in the same sample whereas in two-sample MR the SNP-exposure and the SNP-outcome estimates are obtained from independent samples. In the absence of offspring genotype, paternal genotype is used as a proxy.

As the SNP-exposure association aims to capture the effect of offspring genotype on offspring exposure, it is possible to use the association between own genotype and own exposure from general GWAS summary statistics (assuming that there are no maternal/paternal effects at the locus). Conversely, the SNP-outcome association needs to be obtained from the spousal pairs under study i.e. the association between paternal genetic variants and the maternal outcome. When estimated using paternal genotype, the resulting SNP-outcome association is equal to half that would be estimated using offspring genotype i.e. 0.5 × β^_zx_ × β^_xy_. Consequently, a correction (multiplication by two) needs to be applied to obtain an estimate of the causal effect of the offspring exposure i.e. 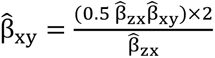. The corresponding standard error can be estimated using the first order terms of the delta method i.e. 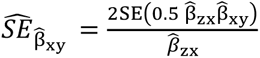. Similarly, this can be extended for the case where causal effects are estimating using more than one SNP (i.e. using inverse variance weighted estimator), by summing the estimated causal effect for each SNP, weighted by the inverse of the variance of the individual causal effect estimates. The use of two-sample MR may improve accuracy of causal estimates by leveraging larger sample sizes for the SNP-exposure association and mitigating any effect of weak instrument bias (Bowden et al., 2019). In addition, the robustness of causal estimates can be investigated using MR sensitivity approaches such as weighted median, weighted mode and MR egger, the latter of which is not appropriate with overlapping samples; (Minelli et al., 2021).

## Conclusion

We have developed a MR approach framework that can be used to estimate the causal effect of offspring traits on parental health outcomes in the absence of offspring genotype data. As there is a dearth of large cohorts of genotyped mother–(or father–) offspring pairs/parent-offspring trios, our method offers an opportunity to provide complementary information on the existence of offspring causal effects owing to the growing availability of large numbers of genome-wide genotyped spousal pairs in publicly available genetic datasets such as the UK Biobank as well as large population based biobanks with linked health record data for first-degree relatives such as HUNT and FinnGen. Whilst we have focused on perinatal offspring traits, this method can also be used to assess the potential effects of postnatal childhood and adult offspring exposures on parental outcomes.

## Contributors

AAH and DME conceived and designed the study. AAH and CBN curated the data. AAH analysed the data and wrote the original draft. AAH, CBN, DAL and DME interpreted the results. All authors critically reviewed the manuscript and approved the final version of the manuscript.

## Data availability

Human genotype and phenotype data from the UKB on which the results of this study were based were accessed with accession ID 53641. The genotype and phenotype data are available upon application to the UKB [http://www.ukbiobank.ac.uk/].

## Ethics

This project received ethical approval from the Institutional Human Research Ethics committee, University of Queensland (Approval Number 2019002705).

