## Supplementary Material for "Utilising offspring genotype by proxy Mendelian randomization to investigate the causal effect of offspring traits on parental health"

#### Supplementary Figures

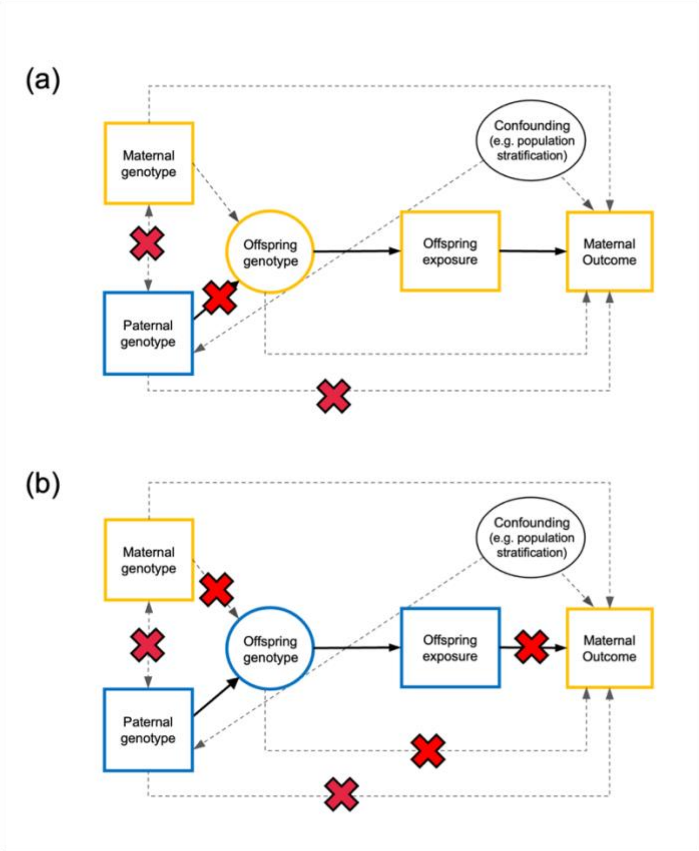

Figure S1: Causal diagrams illustrating spousal pair misclassification and its potential impact on offspring genotype by proxy MR analyses. Within the offspring genotype by proxy MR model, we investigated two misclassification scenarios: (a) A genetically unrelated male is incorrectly used to proxy offspring exposure. Consequently, there is no association between "paternal" genetic variants and the offspring trait and therefore no association between paternal genetic variants and maternal outcome that is mediated through the offspring genome. This could occur when mothers report their offspring's exposure and their own health outcome, but genetic variants from an unrelated male are incorrectly used to instrument her offspring's exposure; (b) A genetically unrelated female is incorrectly paired with the father and his offspring. This could occur when the father reports the offspring trait, and a non-spousal female reports her own health outcome.

### Supplementary Methods

#### *Assumptions of Mendelian Randomization studies of the causal effect of offspring traits on parental health outcomes*

MR relies on a number of assumptions which have previously been discussed at length (Evans & Davey Smith, 2015; Lawlor et al., 2017) (**Error! Reference source not found.**a). First, MR requires that the putative genetic instruments are robustly associated with the exposure of interest in the relevant population (assumption 1 - relevance). Second, MR assumes, not unrealistically given Mendel's Laws of Segregation and Independent Assortment, that these same genetic variants are also uncorrelated with factors that affect the outcome (assumption 2 - independence). Whilst empirical studies suggest that genetic and environmental factors are likely to be independent (Smith et al., 2007), MR analyses that utilise multiple independent SNPs from different genomic regions as instrumental variables (IVs) are potentially more susceptible to violations of this assumption (Yang et al., 2022), in particular owing to unmodelled population structure and familial effects (Brumpton et al., 2020). Third, MR assumes that the genetic variants used are only associated with the outcome through the exposure of interest and not through any alternative independent pathways (assumption 3 – exclusion restriction). This last assumption is particularly problematic for the validity of MR studies and will be violated in the presence of horizontal genetic pleiotropy. This occurs when single nuclear polymorphisms (SNPs; or other genetic polymorphisms in linkage disequilibrium with them) that are associated with the exposure of interest are also associated with the outcome through pathways other than through the exposure. Given the ubiquity of pleiotropy in the human genome, this assumption is likely to be violated in many MR studies (Evans et al., 2013). Variations of the original MR approach that are either robust to some forms of pleiotropy (E.g. MR Egger (Bowden et al., 2015), weighted median, (Bowden et al., 2016), and weighted mode approaches (Hartwig et al., 2017)) and/or permit its detection (Bowden et al., 2018)) have been developed.

MR also relies on the principle of gene-environment equivalence for valid causal inference. This is the notion that genetic perturbations in the exposure have the same effect on the outcome as changes in the exposure induced by environmental factors (Sanderson et al., 2022). These assumptions are necessary for instrument validity and are sufficient to test for the presence of a causal effect of the exposure on the outcome (Didelez & Sheehan, 2007). Additional assumptions are required for estimation of the magnitude of the causal effect, for example, homogeneity of the effect of the exposure on the outcome or monotonicity in the relationship between the genetic variants and the exposure (Richmond & Davey Smith, 2022).

We assume that the genetic instruments act additively on the exposure of interest, with no influence of genetic dominance and epistasis. MR also relies on the assumption that genetic variants in the father are associated with the exposure of interest in the offspring during the period of interest and that this time period is relevant in terms of influencing the maternal outcome. For example, when using birth weight variants as a proxy for fetal growth, we assume these variants capture the influence of fetal growth on the mother during pregnancy.

##### *Assumptions of offspring genotype by proxy MR*

The offspring genotype by proxy MR approach involves the same three core assumptions as conventional MR analyses, however, under this approach there are additional nuances that need to be borne in mind (**Error! Reference source not found.**). Such assumptions are discussed in detail below.

##### Relevance assumption

In the offspring genotype by proxy MR approach, the relevance assumption critically depends upon the accuracy of spousal matching and on the statistical strength of association between offspring genotype and offspring exposure (**Error! Reference source not found.**a). Identification of spousal pairs in the UK Biobank has previously been inferred by matching genotyped, unrelated, opposite sex individuals on the basis of their demographic information (Howe et al., 2019; Rawlik et al., 2019; Robinson et al., 2017; Sjaarda & Kutalik, 2023; Tenesa et al., 2016; Yengo et al., 2018). However, since spousal matching has not been confirmed (e.g. by interview), this classification may be imperfect with some pairs incorrectly identified as spouses (and some spouses failing to be identified), as noted previously (Howe et al., 2019). However, even with extensive demographic information, there is no guarantee that matching of spouses based solely on this knowledge will be perfect. In most biobank style datasets matching would be difficult to definitively confirm without interview or similar verification. It's additionally plausible that within the spousal pairs, an individual's spouse may not be the biological parent of their offspring. For example, in the Avon Longitudinal Study of Parents and Children (ALSPAC), mothers were asked whether their partner was the biological father of their child, with some participants reporting no (Golding et al., 2023), However, the availability of genotyped trios for a subset of the spousal pairs, could provide an indication of how well matching on demographic information alone is likely to perform in the larger cohort(Hatton et al., 2025). For datasets where both parents report their offspring's characteristics, this could provide further reliability. For example, in the UK biobank, both male and female participants report their number of live births/number of children fathered.

For other datasets, spousal matching may be more reliable, particularly when based on large national databases. For example, genotyped individuals in the FinnGen study can be linked to FinRegistry (Viippola et al., 2023) which contains a multigenerational register includes familial relations for first-degree relatives (mother, father, children and siblings). Similarly, in the Nord-Trøndelag Health Study (the HUNT Study), information about married and cohabitating couples is provided by Statistics Norway (Bjørngaard et al., 2017). In other cohorts, such as the Health and Retirement Study (HRS), spousal pairs were established during recruitment, where if the individual was part of a couple, their spouse or partner was also included in the sample (Sonnega et al., 2014).

There will of course be a proportion of spouses who do not have offspring (and therefore offspring phenotype cannot influence maternal outcome by definition) and these pairs should not be included in the main MR analysis. However, if these pairs are correctly identified, they do offer opportunities for sensitivity analyses and identification of likely IV assumption violations (see section below). Finally, confirming that the variance explained in the offspring exposure (assuming offspring phenotype is available) by the paternal genetic variant is roughly what is expected under quantitative genetics theory (i.e. one quarter of the variance of that explained by the regression of own phenotype on own genotype) can also provide an indication of the reliability of the matched data. Relevance could be compromised and/or results biased if the parent being used to proxy offspring genotype is not the biological parent of the offspring in question. The modelling implications of incorrectly matched spousal pairs (including the case where the parent being used to proxy the offspring genetic instrument is not the biological parent) are discussed in the ensuing section on misclassification of spousal pairs.

#### Independence assumption

The independence assumption in MR requires no confounding between the genetic variants and outcomes of interest (**Error! Reference source not found.**b). In standard MR, indirect genetic effects, assortative mating (in previous generations) and population stratification violate this assumption and potentially lead to inconsistent estimates of the causal effect (Davies et al., 2019; Hartwig et al., 2018). When investigating the potential causal effect of offspring traits on their mother's (or father's) health using MR, another potential source of confounding is the maternal (or paternal) genome. This is because maternal genotype will be correlated with offspring genotype (through transmission) and may also plausibly influence the maternal outcome. In the offspring genotype by proxy MR approach, we utilise the paternal genotype to proxy offspring genotype which should be uncorrelated with maternal genotype in the absence of assortative mating and therefore protect MR analyses from confounding by the maternal genome. This relies on the assumption of random mating for the exposure.

There is evidence that spouses typically exhibit greater similarity than would be expected by chance for a variety of traits including height, BMI and education levels. It appears that in many cases this increased similarity is due to phenotypic assortment rather than social homogamy or convergence in phenotypes over time (Robinson et al., 2017). Assortative mating between spousal pairs induces a correlation between spousal genotypes at loci on which the assortment is based (path 2b in **Error! Reference source not found.**b). Thus, phenotypic assortment has the potential to bias causal estimates in the offspring genotype by proxy MR design if the trait undergoing assortment is the exposure of interest or is genetically correlated with (or causes) the exposure. Such assortment can induce correlation between the genetic instruments (or variants in LD with them) in the parents. This potentially opens a path between genetic variants (or variants in LD with them) in the father and the outcome in the mother if the genetic variants for the exposure in the mother are associated with the outcome (**Error! Reference source not found.**b path 3b). The presence of some forms of assortative mating on a single trait could be detected empirically by estimating the correlation between genetic predictors of the trait from SNPs on odd-versus even-numbered chromosomes (Yengo et al., 2018). Assortment is more likely to be problematic when the offspring exposure is a 'visible' trait such as a behavioural phenotype (e.g. educational attainment) or an anthropometric trait (e.g. height and BMI). All these traits have been shown to be subject to assortative mating previously (Robinson et al., 2017; Yengo et al., 2018). In contrast, molecular phenotypes such as lipids and more biologically proximal traits may be less affected by assortment.

#### Exclusion restriction assumption

The exclusion restriction assumption stipulates the genetic instrument must only be associated with the outcome through the exposure. In the current context, it is therefore assumed that there is no directed path between paternal genetic variants and maternal outcome other than that going through the offspring exposure (**Error! Reference source not found.**c). In traditional MR analyses, the exclusion restriction may be violated by horizontal genetic pleiotropy, a feature of the human genome that is likely to be common if not endemic (Evans et al., 2013). This is also the case in offspring genotype by proxy MR where pleiotropic paths from offspring genotype to maternal outcome may also invalidate the use of the paternal genetic variants as instruments (Figure 2c path 1c). However, the offspring genotype by proxy MR framework also has an additional complication, namely the possibility that paternal genetic variants may affect the maternal outcome through the paternal phenotype (**Error! Reference source not found.** path 2c). For example, this assumption would be violated if the exposure of interest, when present in the father were causal for the maternal outcome. While it may be reasonable to assume no causal impact of paternal phenotype on the maternal outcome for some exposures (e.g. if the interest were on the effect of offspring birthweight on maternal health, it is unlikely that paternal birthweight could affect maternal health), this may not be the case for many other exposures. We hypothesise this may be more problematic when the exposure is a behavioural or lifestyle trait whereas molecular phenotypes and perinatal traits may be less problematic. While we recognise that violation of this path is a limitation in the application of offspring genotype by proxy MR to offspring traits more broadly, we discuss the context in which we expect this to be less of a concern in a subsequent section of the manuscript.

Another potential violation of the exclusion restriction assumption is if the paternal genetic variants are pleiotropic for other paternal phenotypes (that are not the proxied exposure) that in turn affect the maternal outcome (Figure 2c path 2c). Potential violations of this assumption could be investigated by examining whether the genetic variants are associated with other paternal phenotypes likely to affect the maternal outcome, as well as employing sensitivity analyses within the offspring genotype by proxy MR context (e.g. MR Egger (Bowden et al., 2015), weighted median, (Bowden et al., 2016), weighted mode (Hartwig et al., 2017), heterogeneity statistic (Bowden et al., 2018)). Where violations are identified, it may be possible to perform sub-analyses excluding the potentially pleiotropic variants. Similarly, the presence of horizontal pleiotropy can be investigated by incorporating gene by environment interactions into the offspring genotype by proxy MR approach as discussed in a subsequent section (Davey Smith, 2011; Spiller et al., 2019). While dynastic effects of paternal genotype on offspring exposure (path 3c) do not violate the exclusion restriction assumption, these must be accounted for when estimating the causal effect.

#### Additional considerations - Indirect genetic effects on the offspring exposure

The offspring genotype by proxy MR approach, leverages the assumption that the genetic variants for the exposure in one parent exert their effect on the offspring exposure through the offspring's genome (i.e. following transmission of alleles from parent to offspring). However, these same parental genotypes may also exert indirect (or dynastic) effects on offspring exposures (Hwang et al., 2021; Kong et al., 2018; Warrington et al., 2018). Therefore, it is possible that the parental IV may also (potentially inadvertently) capture parental indirect genetic effects on the offspring exposure. However, provided the parental genetic variants are (strongly) marginally associated with the offspring exposure (i.e. direct offspring and indirect parental genetic effects on the offspring exposure don't cancel out), this fact should not be problematic in and of itself for causal inference, as both parental and offspring genetic variants may constitute valid IVs. We show mathematically that dynastic effects of the paternal genotype on the offspring exposure does not result in inconsistent estimates of the causal effect in the offspring genotype by proxy MR design in the supplementary material. However, parental genetic variants that exert indirect effects on some offspring exposures may also be more likely to exert effects on other offspring phenotypes and/or spousal phenotypes (see discussion of the exclusion restriction assumption above). For example, genetic variants that are associated with alcohol consumption in fathers, could conceivably affect a range of offspring and maternal phenotypes through pathways other than through offspring alcohol consumption. We hypothesise that these issues may be more problematic when the exposure of interest is an overtly observable phenotype (e.g. education attainment, smoking, alcohol consumption, diet, physical activity etc), as opposed to more biologically proximal traits (such as a circulating biomarker).

##### *Inconsistency of instrumental variable estimators under pleiotropy*

The large sample properties of each instrumental variable estimator can be derived under asymptotic theory. For the subsequent derivations, we define the following quantities:

$Z_p$ = the genetic variant in the fathers

$Z_m$ = the genetic variant in the mothers

$Z_o$ = the genetic variant in the offspring

$Z_{pt}$ = the allele transmitted from father to offspring

X = offspring exposure

Y = maternal outcome

$\beta_{ZO,X}$ = population level association between offspring genetic variant and offspring exposure

$\beta_{XY}$ = population level causal effect of offspring exposure on maternal outcome

$\beta_{ZO,Y}$ = population level effect of offspring genetic variant on maternal outcome

$\beta_{ZP,Y}$ = population level effect of paternal genetic variant on maternal outcome

$\beta_{ZP,X}$ = population level effect of paternal genetic variant on offspring exposure

For all derivations, we assume absence of assortative mating and genetic confounding through the maternal genome.

In the case of the offspring genotype by proxy MR design, the causal effect in large samples is given by:

$$\text{plim}(\hat{\beta}_{IV}) = \text{plim}\left(\frac{\text{cov}(Z_P,Y)}{\text{cov}(Z_P,X)}\right) = \frac{\text{COV}(Z_P,Y)}{\text{COV}(Z_P,X)} = \frac{(\beta_{ZP,Y}+0.5\times\beta_{ZO,Y}+0.5\times\beta_{XY}\times\beta_{ZO,X})\times\text{var}(Z_P)}{0.5\times\beta_{ZO,X}\times\text{var}(Z_P)} = \beta_{XY} + \frac{\beta_{ZO,Y}}{\beta_{ZO,X}} + \frac{2\beta_{ZP,Y}}{\beta_{ZO,X}}$$

where plim is the probability limit, cov refers to the sample covariance, and COV and VAR the covariance and variance in the population respectively.

In the case of MR with adjustment for maternal genotype, the causal effect in large samples is given by:

$$\text{plim}(\hat{\beta}_{IV}) = \text{plim}\left(\frac{\text{cov}(Z_O,Y)}{\text{cov}(Z_O,X)}\right) = \frac{\text{COV}(Z_O,Y)}{\text{COV}(Z_O,X)} = \frac{\beta_{XY}\times\beta_{ZO,X}\times\text{var}(Z_O)+0.5\times\beta_{ZP,Y}\times\text{var}(Z_P)+\beta_{ZO,Y}\times\text{var}(Z_O)}{\beta_{ZO,X}\times\text{var}(Z_O)} = \beta_{XY} + \frac{\beta_{ZO,Y}}{\beta_{ZO,X}} + \frac{\beta_{ZP,Y}}{2\beta_{ZO,X}}$$

In the case of MR using paternally transmitted alleles, the causal effect in large samples is given by:

$$\text{plim}(\hat{\beta}_{IV}) = \text{plim}\left(\frac{\text{cov}(Z_{Pt},Y)}{\text{cov}(Z_{Pt},X)}\right) = \frac{\text{COV}(Z_{Pt},Y)}{\text{COV}(Z_{Pt},X)}$$

$$= \frac{\beta_{XY}\times\beta_{ZO,X}\times\frac{1}{2}\times\text{var}(Z_O)+\beta_{ZO,Y}\times\frac{1}{2}\times\text{var}(Z_O)+\beta_{ZP,Y}\times\frac{1}{2}\times\text{var}(Z_O)}{\beta_{ZO,X}\times\frac{1}{2}\times\text{var}(Z_O)}$$

$$= \beta_{XY} + \frac{\beta_{ZO,Y}}{\beta_{ZO,X}} + \frac{\beta_{ZP,Y}}{\beta_{ZO,X}}$$

For the last derivation, we assume that the parental origins of the alleles transmitted to the offspring can be determined with certainty.

Finally, we show that direct effects of the paternal genotype on the offspring exposure does not result in inconsistent estimates of the causal effect in the offspring genotype by proxy MR design:

$$\text{plim}(\hat{\beta}_{IV}) = \text{plim}\left(\frac{\text{cov}(Z_P,Y)}{\text{cov}(Z_P,X)}\right) = \frac{\text{COV}(Z_P,Y)}{\text{COV}(Z_P,X)}$$

$$= \frac{0.5\times\beta_{XY}\times\beta_{ZO,X}\times\text{var}(Z_P)+\beta_{XY}\times\beta_{ZP,X}\times\text{var}(Z_P)}{0.5\times\beta_{ZO,X}\times\text{var}(Z_P)+\beta_{ZP,X}\times\text{var}(Z_P)} = \beta_{XY}$$

##### *Methods for data simulations*

In this section we describe the methods used for data simulation. In all simulations we use the illustrative example of a potential causal effect of an offspring exposure on their mother's health. Data simulations include:

(1) To explore the likely consequences of spousal pair misclassification
(2) To estimate power to detect the causal effect of an offspring exposure on a maternal health outcome using MR

For each simulation, we generated maternal, paternal and offspring genotypes at a single genetic locus (even if not all of these were used in the analysis). Genotypes were simulated assuming a trait increasing allele frequency of q = 0.6 and standard autosomal Mendelian inheritance. Standardized additive genotypic dosages (mean zero and unit variance) for maternal ($Z_m$), paternal ($Z_p$) and offspring ($Z_o$) genotypes were calculated. For each family i, the offspring exposure X was generated using the following equation:

$$X_i = \sqrt{V_q}\times Z_{o_i} + \beta_{UX}\times U_i + \delta_i$$

where $V_q$ denotes the variance in the offspring exposure explained by offspring genotype, $Z_o$ is a latent variable of unit variance indexing the offspring genotype, U is a standard normal random variable representing (unmeasured) confounding influences, $\beta_{UX}$ denotes the total effect of latent confounders U on the offspring exposure X, and δ is a normally distributed random variable with mean zero. The variance of δ is such that X has unit variance.

The maternal outcome Y (for each family i) was generated according to the following model:

$$Y_i = \beta_{XY} \times X_i + \beta_{UY} \times U_i + \sqrt{V_o} \times Z_{o_i} + \sqrt{V_f} \times Z_{p_i} + \sqrt{V_m} \times Z_{m_i} + \epsilon_i$$

where $\beta_{XY}$ is the causal effect of the offspring exposure X on the maternal outcome Y, $\beta_{UY}$ is the total effect of confounding variables on the maternal outcome, $V_o$ denotes the variance in the maternal outcome explained by offspring genotype, $Z_o$ is a latent variable of unit variance indexing the offspring genetic instrument, $V_p$ denotes the variance in the maternal outcome explained by paternal genotype, $Z_p$ is a latent variable of unit variance indexing the paternal genetic instrument, $V_m$ denotes the variance in the maternal outcome explained by maternal genotype, $Z_m$ is a latent variable of unit variance indexing the maternal genetic instrument, and $\epsilon$ is a random normal variate with mean zero with variance such that Y has unit variance asymptotically.

$\beta_{XY}$ represents the causal effect of the offspring exposure X on the maternal outcome Y that we estimate from the MR analyses. However, we know (as shown in the Figure 1d) that this will likely be biased because in the presence of non-zero paths from the offspring genetic instrument $Z_o$ and the paternal genetic instrument $Z_p$ to the maternal outcome Y.

#### Methods for misclassification of spousal pairs

We considered the consequences of (accidental) inclusion of incorrectly matched spousal pairs (or correctly matched spousal pairs that are not biological parents) on bias and statistical power. We performed a simple simulation study to identify the likely effect of such misclassification varying the proportion of randomly misclassified spouses (0%, 10%, 20%, 30% and 40% of spousal pairs), utilising the illustrative example of potential causal effects of offspring perinatal exposures on maternal post-natal health. Within this model we investigated two misclassification scenarios; (1) A male who is genetically unrelated to the offspring under consideration is incorrectly used to the proxy offspring exposure. Consequently, there is no association between paternal genetic variants, offspring genotype and offspring exposure and therefore no association between paternal genetic variants and maternal outcome through this pathway. This could occur when mothers report their offspring's exposure and their own health outcome, but genetic variants from an unrelated male are incorrectly used to instrument her offspring's exposure; (2) A genetically unrelated female is incorrectly paired with the father and his offspring. This could occur when the father reports the offspring trait, and a non-spousal female reports her own health outcome.

In the main simulations, we considered a true causal effect of $\beta_{XY}$ = 0.2 and a sample size of 50,000 spousal pairs (which corresponds to the approximate number of spousal pairs in the UK Biobank). Offspring genotype was simulated to explain 1%, 2% or 5% of the variance in the offspring exposure, which corresponded to paternal genotype explaining 0.25%, 0.5% or 1.25% of the variance, respectively. MR analyses were performed using two-stage least squares using the ivreg R package version 0.6-5 (John Fox, 2024). For each scenario, we generated 1,000 replicates and recorded the median instrumental variables estimate of the causal relationship, it's standard error and the power/type 1 error rate of the analysis. Full simulation results in supplementary table 1 where we consider a true causal effect of $\beta_{XY}$ = 0.2, 0 and -0.2.

#### Methods for power of the offspring genotype by proxy MR approach

We investigated the power of three different MR approaches: (1) Offspring genotype by proxy MR (Figure 1d), (2) MR with adjustment for maternal genotype (Figure 1b) and (3) MR using paternally transmitted alleles (Figure 1c). Data were generated following the data simulation procedure outlined above. For power calculations, simulations were used to estimate the sample sizes required to detect a standardized causal effect of $\beta_{XY}$ = 0.1 at 80% power (assuming Type 1 error rate of $\alpha$ = 0.05) using the mean of 1,000 replicates. The sample size refers to the relevant observational unit as per the study design i.e. (1) a spousal pair, (2) a mother-offspring pair, (3) a parent offspring trio. Sample sizes ranging from 7,000 to 300,000 units were used for power calculations. We also varied the size of the effect of offspring genotype on offspring exposure ($V_o$) ranging from 0.01 to 0.1 and set the strength of confounding between the offspring exposure and maternal outcome at 0.5. MR analyses were performed using two-stage least squares using the ivreg R package version 0.6-5 (John Fox, 2024). For each scenario, we generated 1,000 replicates and recorded the median instrumental variables estimate of the causal relationship, it's standard error and the power/type 1 error rate of the analysis. For power calculations we do not consider the potential effects of pleiotropy (i.e. variant in the maternal outcome explained by offspring ($\mathrm{V_o}$), maternal ($\mathrm{V_m}$) and paternal ($\mathrm{V}_p$) genotype).

### Supplementary Tables

Supplementary Table 1: Results from the simulation study on the impact of spousal pair misclassification

Supplementary Table 2: Results from power calculations by data simulation and asymptotic theory
